# Higher dose corticosteroids in hospitalised COVID-19 patients requiring ventilatory support (RECOVERY): a randomised, controlled, open-label, platform trial

**DOI:** 10.1101/2024.09.04.24312992

**Authors:** RECOVERY Collaborative Group, Peter W Horby, Jonathan R Emberson, Louise Thwaites, Mark Campbell, Leon Peto, Guilherme Pessoa-Amorim, Natalie Staplin, Raph L Hamers, John Amuasi, Jeremy Nel, Evelyne Kestelyn, Nguyen Thanh Phong, Anil Shrestha, Nasronudin Nasronudin, Rahuldeb Sarkar, Pham Ngoc Thach, Damodar Patel, Uun Samardi, Richard Stewart, Erni Nelwan, Manisha Rawal, J Kenneth Baillie, Maya H Buch, Jeremy N Day, Saul N Faust, Thomas Jaki, Katie Jeffery, Edmund Juszczak, Marian Knight, Wei Shen Lim, Marion Mafham, Alan Montgomery, Andrew Mumford, Kathryn Rowan, Buddha Basnyat, Richard Haynes, Martin J Landray

**Author notes:** Correspondence to: Prof Peter W Horby and Prof Martin J Landray, RECOVERY Central Coordinating Office, Richard Doll Building, Old Road Campus, Roosevelt Drive, Oxford OX3 7LF, United Kingdom. The Writing Committee and Trial Steering Committee are listed at the end of this manuscript and a complete list of collaborators in the Randomised Evaluation of COVID-19 Therapy (RECOVERY) trial is provided in the Supplementary Appendix.

## Abstract

**Background:** Low-dose corticosteroids (e.g. 6 mg dexamethasone) have been shown to reduce mortality for hypoxic COVID-19 patients. We have previously reported that higher dose corticosteroids cause harm in patients with hypoxia but not receiving ventilatory support (non-invasive mechanical ventilation, invasive mechanical ventilation or extra-corporeal membrane oxygenation), but the balance of efficacy and safety in patients receiving ventilatory support is uncertain.

**Methods:** This randomised, controlled, open-label platform trial (Randomised Evaluation of COVID-19 Therapy [RECOVERY]) assessed multiple possible treatments in patients hospitalised for COVID-19. Eligible and consenting adult patients receiving ventilatory support were randomly allocated (1:1) to either usual care with higher dose corticosteroids (dexamethasone 20 mg once daily for 5 days followed by 10 mg once daily for 5 days or until discharge if sooner) or usual standard of care alone (which includes dexamethasone 6 mg once daily for 10 days or until discharge if sooner). The primary outcome was 28-day mortality; secondary outcomes were duration of hospitalisation and (among participants not on invasive mechanical ventilation at baseline) the composite of invasive mechanical ventilation or death. Recruitment closed on 31 March 2024 when funding for the trial ended. The RECOVERY trial is registered with ISRCTN (50189673) and clinicaltrials.gov (NCT04381936).

**Findings:** Between 25 May 2021 and 9 January 2024, 477 COVID-19 patients receiving ventilatory support were randomly allocated to receive usual care plus higher dose corticosteroids versus usual care alone (of whom 99% received corticosteroids during the follow-up period). Of those randomised, 221 (46%) were in Asia, 245 (51%) in the UK and 11 (2%) in Africa. 143 (30%) had diabetes mellitus. Overall, 86 (35%) of 246 patients allocated to higher dose corticosteroids versus 86 (37%) of 231 patients allocated to usual care died within 28 days (rate ratio [RR] 0.87; 95% CI 0.64-1.18; p=0.37). There was no significant difference in the proportion of patients discharged from hospital alive within 28 days (128 [52%] in the higher dose corticosteroids group vs 120 [52%] in the usual care group; RR 1.04, 0.81-1.33]; p=0.78). Among those not on invasive mechanical ventilation at baseline, there was no significant difference in the proportion meeting the composite endpoint of invasive mechanical ventilation or death (76 [37%] of 206 vs 93 [45%] of 205; RR 0.79 [95% CI 0.63–1.00]; p=0.05).

**Interpretation:** In patients hospitalised for COVID-19 receiving ventilatory support, we found no evidence that higher dose corticosteroids reduced the risk of death compared to usual care, which included low dose corticosteroids.

**Funding:** UK Research and Innovation (Medical Research Council) and National Institute of Health Research (Grant ref: MC_PC_19056), and Wellcome Trust (Grant Ref: 222406/Z/20/Z).

## INTRODUCTION

The RECOVERY trial has previously shown that the use of corticosteroids (using dexamethasone 6 mg once daily for ten days or until discharge if sooner) reduces the risk of death in patients admitted to hospital with COVID-19 and hypoxia.^1^ Subsequent findings that additional immunosuppression with an interleukin-6 (IL-6) receptor blocker and/or a Janus kinase (JAK) inhibitor further reduces the risk of death in these patients raised the question whether simply increasing the dose of corticosteroid rather than adding other immunomodulators could confer additional benefits at substantially lower cost.^2,3^

In April 2021, the United Kingdom COVID-19 Therapeutics Advisory Panel recommended that the RECOVERY trial study higher dose of corticosteroids.^4^ The RECOVERY trial therefore established a randomised evaluation of the effects of higher-dose corticosteroids versus usual care for adult patients who had been admitted to hospital with COVID-19 and had clinical evidence of hypoxia. Usual care for hypoxic COVID-19 patients includes low dose corticosteroids. On 11 May 2022, the independent Data Monitoring Committee recommended that this comparison be halted for those patients receiving no oxygen or simple oxygen only on the grounds of safety and the results among such patients were published, showing that higher dose corticosteroids were associated with an increased 28-day mortality (123/659 [19%] in those allocated higher dose corticosteroids versus 75/613 [12%] in those allocated usual care, rate ratio 1.59; 95% confidence interval [CI] 1.20-2.10; p=0.0012.^5^ Recruitment of patients receiving ventilatory support continued.

Trials of corticosteroids in non-COVID acute respiratory distress syndrome (ARDS) have not directly compared higher doses with the lower dose used in RECOVERY.^6^ Higher doses of corticosteroids have been assessed in other trials among patients with COVID-19 receiving ventilatory support, but none have demonstrated clear benefit or harm.^7–11^ A WHO meta-analysis of randomised controlled trials in critically ill COVID-19 indicated similar mortality benefit with lower and higher-dose corticosteroids, but estimates were imprecise.^12^

Here we report the results of the randomised comparison from the RECOVERY trial comparing higher dose corticosteroids with usual care among patients receiving ventilatory support.

## METHODS

### Study design and participants

The Randomised Evaluation of COVID-19 therapy (RECOVERY) trial is an investigator-initiated, individually randomised, controlled, open-label, platform trial to evaluate the effects of potential treatments in patients hospitalised with COVID-19. Details of the trial design and results for other possible treatments (dexamethasone, hydroxychloroquine, lopinavir-ritonavir, azithromycin, tocilizumab, convalescent plasma, colchicine, aspirin, casirivimab plus imdevimab, baricitinib, higher-dose corticosteroids among patients not receiving ventilatory support, empagliflozin, molnupiravir and nirmatrelvir-ritonavir) have been published previously.^1–3,5,13–21^ The trial was conducted at hospital organisations in the United Kingdom, supported by the National Institute for Health Research Clinical Research Network, as well as in South and Southeast Asia and Africa. Of these, 81 hospitals in the UK, 5 in Nepal, 2 in Indonesia, 2 in Vietnam, 2 in South Africa, and 1 in Ghana enrolled participants in the evaluation of higher dose corticosteroids (appendix pp 5-31). The trial was coordinated by the Nuffield Department of Population Health at the University of Oxford (Oxford, UK), the trial sponsor. The trial was conducted in accordance with the principles of the International Conference on Harmonisation–Good Clinical Practice guidelines. The protocol was approved by all relevant regulatory authorities and ethics committees in each participating country (appendix p 32) The protocol and statistical analysis plan are available in the appendix (pp 69-151) with additional information available on the study website www.recoverytrial.net.

Patients aged at least 18 years admitted to hospital were eligible for the study if they had clinically suspected or laboratory confirmed SARS-CoV-2 infection, clinical evidence of hypoxia (i.e. receiving oxygen with or without other forms of respiratory support, or with oxygen saturations <92% on room air) and no medical history that might, in the opinion of the attending clinician, put the patient at significant risk if they were to participate in the trial. Patients were ineligible for the comparison of higher dose corticosteroid vs. usual care if there was a known contra-indication to short-term use of corticosteroids, suspected or confirmed influenza, or current use of nirmatrelvir-ritonavir, ritonavir or other potent CYP3A inhibitors. Ventilatory support includes continuous positive airway pressure, bi-level positive airway pressure, high-flow nasal oxygen (together non-invasive ventilation), invasive mechanical ventilation and extracorporeal membrane oxygenation (ECMO). Endemic infections were screened for in accordance with local practice. Other immunomodulatory therapies were not contra-indicated but investigators were advised to consider the total burden of such therapy (e.g. combining an IL-6 receptor antagonist with higher dose corticosteroid). Written informed consent was obtained from all patients, or a legal representative if patients were too unwell or unable to provide consent.

### Randomisation and masking

Baseline data were collected using a web-based case report form that included demographics, level of respiratory support, major comorbidities, suitability of the study treatment for a particular patient, SARS-CoV-2 vaccination status, and treatment availability at the study site (appendix pp 46).

Eligible and consenting patients were assigned in a 1:1 ratio to either usual standard of care plus higher dose corticosteroids or usual standard of care alone (which includes low dose corticosteroids, usually dexamethasone 6 mg once daily for 10 days or until discharge if sooner), using web-based simple (unstratified) randomisation with allocation concealed until after randomisation (appendix pp 41-5).^22,23^ Patients allocated to higher dose corticosteroid were to receive dexamethasone 20 mg daily for 5 days followed by dexamethasone 10 mg for 5 days (or until discharge if sooner). Alternative corticosteroid regimens for pregnant women are described in the protocol (appendix p 69-111).

As a platform trial, and in a factorial design, patients could be simultaneously randomised to other treatment groups: i) empagliflozin versus usual care, ii) sotrovimab versus usual care, and iii) molnupiravir versus usual care. Further details of when these factorial randomisations were open are provided in the supplementary appendix (pp 41-3). Participants and local study staff were not masked to the allocated treatment. Other than members of the Data Monitoring Committee, all individuals involved in the trial were masked to aggregated outcome data while recruitment and 28-day follow-up were ongoing.

### Procedures

An online follow-up form was completed by site staff when patients were discharged, had died, or at 28 days after randomisation, whichever occurred first (appendix pp 47-55). Information was recorded on adherence to allocated trial treatment, receipt of other COVID-19 treatments, duration of admission, receipt of respiratory or renal support, new cardiac arrhythmia, thrombosis, clinically significant bleeding, non-COVID infection, metabolic complications (collected from 28 July 2021 onwards), and vital status (including cause of death). In addition, in the UK, routinely collected healthcare and registry data were obtained, including information on vital status at day 28 (with date and cause of death); discharge from hospital; and receipt of respiratory support or renal replacement therapy. For sites outside the UK a further case report form (appendix p 56) collected vital status at day 28 (if not already reported on follow-up form).

### Outcomes

Outcomes were assessed at 28 days after randomisation, with further analyses specified at 6 months. The primary outcome was 28-day all-cause mortality. Secondary outcomes were time to discharge from hospital, and, among patients not on invasive mechanical ventilation at randomisation (which for this report means those who are on non-invasive mechanical ventilation at randomisation), the composite outcome of invasive mechanical ventilation (including extra-corporeal membrane oxygenation) or death. Prespecified subsidiary clinical outcomes were successful cessation of invasive ventilation among those receiving invasive ventilation at randomisation, use of invasive mechanical ventilation among patients not on non-invasive ventilation at randomisation, and use of renal dialysis or haemofiltration. Prespecified safety outcomes were cause-specific mortality, major cardiac arrhythmia, thrombotic and major bleeding events, other infections and metabolic complications. Information on suspected serious adverse reactions was collected in an expedited fashion to comply with regulatory requirements. Details of the methods used to ascertain and derive outcomes are provided in the appendix (pp 152).

### Sample size and role of the independent Data Monitoring Committee

As stated in the protocol, appropriate sample sizes could not be estimated when the trial was being planned. However, the intention for this comparison was to continue recruitment until sufficient primary outcomes had accrued to have 90% power to detect a proportional risk reduction of 20% at 2P=0.01.

The independent Data Monitoring Committee reviewed unblinded analyses of the study data and any other information considered relevant to the trial at intervals of around 2-3 months (depending on speed of enrolment) and was charged with determining if, in their view, the randomised comparisons in the study provided evidence on mortality that was strong enough (with a range of uncertainty around the results that was narrow enough) to affect national and global treatment strategies (appendix p 58).

On 11 May 2022, the Data Monitoring Committee recommended stopping recruitment to the higher dose corticosteroid comparison for patients who require no oxygen or simple oxygen only at randomisation due to safety concerns (appendix p 59). The Data Monitoring Committee encouraged continuing recruitment of all those patients who, at randomisation, require either non-invasive ventilation, invasive mechanical ventilation or ECMO. Recruitment continued until 31 March 2024 when funding for the trial ended.

### Statistical Analysis

All analyses in this report were limited to the subgroup of patients receiving ventilatory support at randomisation. For all outcomes, intention-to-treat analyses compared patients randomised to higher dose corticosteroids with patients randomised to usual care. For the primary outcome of 28-day mortality, the HR from a Cox model with adjustment for age in three categories (<70 years, 70-79 years, and 80 years or older) and ventilation status at randomisation in two categories (non-invasive ventilation and invasive mechanical ventilation) was used to estimate the mortality rate ratio. We constructed Kaplan-Meier survival curves to display cumulative mortality over the 28-day period (starting on the day of randomisation and ending 28 days later). We used the same Cox regression method to analyse time to hospital discharge and successful cessation of invasive mechanical ventilation, with patients who died in hospital right-censored on day 29. Median time to discharge was derived from Kaplan-Meier estimates. For the composite secondary outcome of progression to invasive mechanical ventilation or death within 28 days, and the subsidiary clinical outcomes of receipt of ventilation and use of haemodialysis or haemofiltration, the precise dates were not available and a log-binomial regression model was used to estimate the risk ratio (RR) adjusted for age and ventilation status. Estimates of rate and risk ratios (both denoted RR) are shown with 95% confidence intervals. For safety outcomes, unadjusted absolute risk differences were calculated as the difference in the proportions of patients experiencing outcomes by treatment allocation.

Since the analyses presented here relate only to the subset of participants who required ventilatory support at randomisation, any analyses of the primary outcome in further subgroups defined by different baseline characteristics must be considered exploratory in nature. With that caveat, we present analyses of the primary outcome by age, sex, ethnicity, country, days since symptom onset, and respiratory support received with tests of heterogeneity or trend, as appropriate. We have not presented analyses by subgroups for the secondary or other outcomes. Results for the pre-specified other clinical outcomes and safety outcomes are presented. For the primary outcome of 28-day mortality, the results from RECOVERY were subsequently included in a meta-analysis of results from all previous randomised trials of higher vs lower dose steroids in patients with COVID-19. For each trial, we compared the observed number of deaths among patients allocated higher dose steroids with the expected number if all patients were at equal risk (ie, we calculated the observed minus expected statistic [o–e], and its variance v). For the previously-reported RECOVERY findings in lower risk (not on ventilation) patients, these were taken as the log-rank observed minus expected statistic and its variance but for other trials, where the exact timing of each death was not available, these were calculated from standard formulae for 2 × 2 contingency tables. We then combined trial results using the log of the mortality rate ratio calculated as the inverse-variance weighted average S/V with variance 1/V (and hence with 95% CI S/V ±1.96/√V), where S is the sum over all trials of (O–E) and V is the sum over all trials of v. The full database is held by the study team which collected the data from study sites and performed the analyses at the Nuffield Department of Population Health, University of Oxford (Oxford, UK). Analyses were performed using SAS version 9.4 and R version 4.0.3. The trial is registered with ISRCTN (50189673) and clinicaltrials.gov (NCT04381936).

## RESULTS

Recruitment to the evaluation of higher dose corticosteroids commenced on 25 May 2021 outside the UK and 29 December 2021 in the UK (following closure of the baricitinib comparison) and ended worldwide on 31 March 2024 (last participant randomised on 9 January 2024). Of 1749 patients enrolled in this comparison during this period, 477 patients receiving ventilatory support are included in this evaluation. Of these, 246 were randomly allocated to higher dose corticosteroids and 231 patients were randomly allocated to usual care (Figure 1). The mean age of these participants was 61.4 years (SD 15.5), 221 (46%) were recruited in Asia, 245 (51%) in the UK and 11 (2%) in Africa. 411 (86%) were receiving non-invasive ventilation and 66 (14%) were receiving invasive mechanical ventilation. 143 (30%) had a history of diabetes mellitus (Table 1).

**Figure 1:**
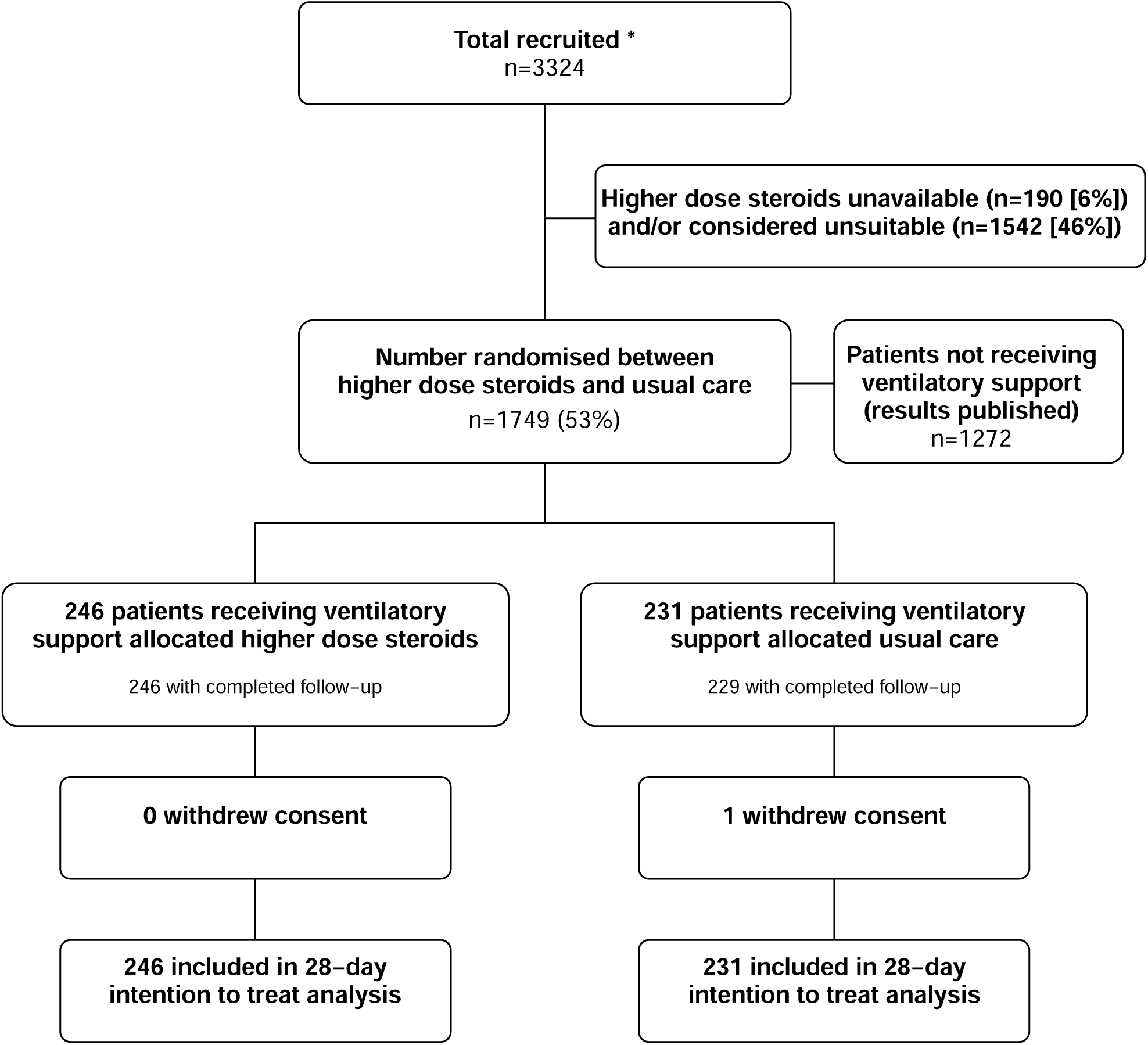
Trial profile. ITT=intention to treat. Higher dose corticosteroid unavailable and higher dose corticosteroid considered unsuitable are not mutually exclusive. High risk patients are those that were receiving non-invasive ventilation, invasive mechanical ventilation or ECMO at randomisation. Low risk patients are those that were hypoxic but receiving no oxygen or simple oxygen only. * Number recruited overall during period that participants on no oxygen or simple oxygen only could be recruited into the higher dose corticosteroid comparison.

**Table 1:** Baseline characteristics by treatment allocation.

|  | Higher dose steroids<br>(n=246) | Usual care<br>(n=231) |
| --- | --- | --- |
| Age, years | 61.7 (14.9) | 61.1 (16.1) |
| <70 | 164 (67%) | 157 (68%) |
| ≥70 to <80 | 61 (25%) | 46 (20%) |
| ≥80 | 21 (9%) | 28 (12%) |
| Sex |  |  |
| Male | 147 (60%) | 151 (65%) |
| Female | 99 (40%) | 80 (35%) |
| Country |  |  |
| Indonesia | 28 (11%) | 37 (16%) |
| Nepal | 44 (18%) | 42 (18%) |
| South Africa | 8 (3%) | 3 (1%) |
| Vietnam | 40 (16%) | 30 (13%) |
| United Kingdom | 126 (51%) | 119 (52%) |
| Ethnicity |  |  |
| White | 107 (43%) | 96 (42%) |
| Black | 1 (<0.5%) | 5 (2%) |
| Asian | 77 (31%) | 87 (38%) |
| Other | 4 (2%) | 3 (1%) |
| Unknown | 57 (23%) | 40 (17%) |
| Number of days since symptom onset | 8 (5-12) | 7 (5-12) |
| Number of days since admission to hospital | 2 (1-4) | 2 (1-4) |
| Respiratory support received |  |  |
| Non-invasive ventilation | 206 (84%) | 205 (89%) |
| Invasive mechanical ventilation | 40 (16%) | 26 (11%) |
| Previous diseases |  |  |
| Diabetes | 74 (30%) | 69 (30%) |
| Heart disease | 77 (31%) | 69 (30%) |
| Chronic lung disease | 57 (23%) | 46 (20%) |
| Tuberculosis | 1 (<0.5%) | 2 (1%) |
| HIV | 4 (2%) | 1 (<0.5%) |
| Severe liver disease * | 7 (3%) | 4 (2%) |
| Severe kidney impairment † | 22 (9%) | 14 (6%) |
| Any of the above | 159 (65%) | 134 (58%) |
| SARS-CoV-2 test result |  |  |
| Positive | 232 (94%) | 213 (92%) |
| Negative | 2 (1%) | 3 (1%) |
| Unknown | 12 (5%) | 15 (6%) |
| Received a COVID-19 vaccine | 133 (54%) | 112 (48%) |
| Use of other treatments |  |  |
| Remdesivir | 88 (36%) | 78 (34%) |
| Tocilizumab | 57 (23%) | 51 (22%) |
| Plan to use tocilizumab within the next 24 hours | 23 (9%) | 17 (7%) |

Data are mean (SD), n (%), or median (IQR). 0 pregnant women were randomised. \* Defined as requiring ongoing specialist care.
† Defined as estimated glomerular filtration rate <30 mL/min per 1.73 m<sup>2</sup>

The follow-up form was completed for 246 (100%) patients in the higher dose corticosteroid group and 229 (99.1%) patients in the usual care group. Among patients with a completed follow-up form, 87% allocated to higher dose corticosteroid were reported to have received higher dose corticosteroids compared with 6% allocated to usual care (figure 1, webtable 2). Among those with a completed follow-up form allocated usual care, 68% received low dose and 18% received intermediate dose (>6 <20 mg) dexamethasone. 39% received remdesivir, 15% received an interleukin-6 antagonist and 5% received baricitnib during the follow-up period (webtable 2).

Primary and secondary outcome data are known for >99% of randomly assigned patients. Allocation to higher dose corticosteroids was associated with a non-significant reduction in the primary outcome of 28-day mortality compared with usual care alone: 86 (35%) of 246 patients in the higher dose corticosteroid group died vs 86 (37%) of 231 patients in the usual care group (rate ratio 0.87; 95% CI 0.64-1.18; p=0.37; table 2, figure 2). In exploratory analyses, there was no good evidence that the proportional effect of higher dose corticosteroids on mortality differed across all 5 pre-specified subgroups, nor in an exploratory analysis by country (Figure 3). In another exploratory analysis (data not shown), there was also no good evidence of heterogeneity according to baseline use (or planned use) of tocilizumab.

**Table 2:** Effect of allocation to higher dose corticosteroid on key study outcomes.

|  | Treatment allocation |  | RR (95% CI) |
| --- | --- | --- | --- |
|  | Higher dose steroids (n=246) | Usual care (n=231) |  |
| **Primary outcome** |  |  |  |
| 28-day mortality | 86 (35%) | 86 (37%) | 0.87 (0.64-1.18) |
| **Secondary outcomes** |  |  |  |
| Time to being discharged alive, days | 24 (12 to >28) | 26 (11 to >28) |  |
| Discharged from hospital within 28 days | 128 (52%) | 120 (52%) | 1.04 (0.81-1.33) |
| Receipt of invasive mechanical ventilation or death* | 76/206 (37%) | 93/205 (45%) | 0.79 (0.63-1.00) |
| Invasive mechanical ventilation | 29/206 (14%) | 40/205 (20%) | 0.72 (0.46-1.11) |
| Death | 65/206 (32%) | 79/205 (39%) | 0.80 (0.62-1.04) |
| **Subsidiary clinical outcomes** |  |  |  |
| Successful cessation of invasive mechanical ventilation † | 14/40 (35%) | 11/26 (42%) | 0.80 (0.36-1.80) |
| Use of haemodialysis or haemofiltration ‡ | 21/243 (9%) | 18/229 (8%) | 0.97 (0.54-1.74) |

Data are n (%) or n/N (%), unless otherwise indicated. RR=rate ratio for the outcomes of 28-day mortality and hospital discharge, and risk ratio for other outcomes. CI=confidence interval. Estimates of the RR and its 95% CI are adjusted for age in three categories (<70 years, 70-79 years, and 80 years or older) and ventilation status at randomisation in two categories (non-invasive ventilation and invasive mechanical ventilation). \* Analyses exclude those on invasive mechanical ventilation at randomisation. † Analyses restricted to those on invasive mechanical ventilation at randomisation. ‡ Analyses exclude those on haemodialysis or haemofiltration at randomisation.

**Figure 2:**
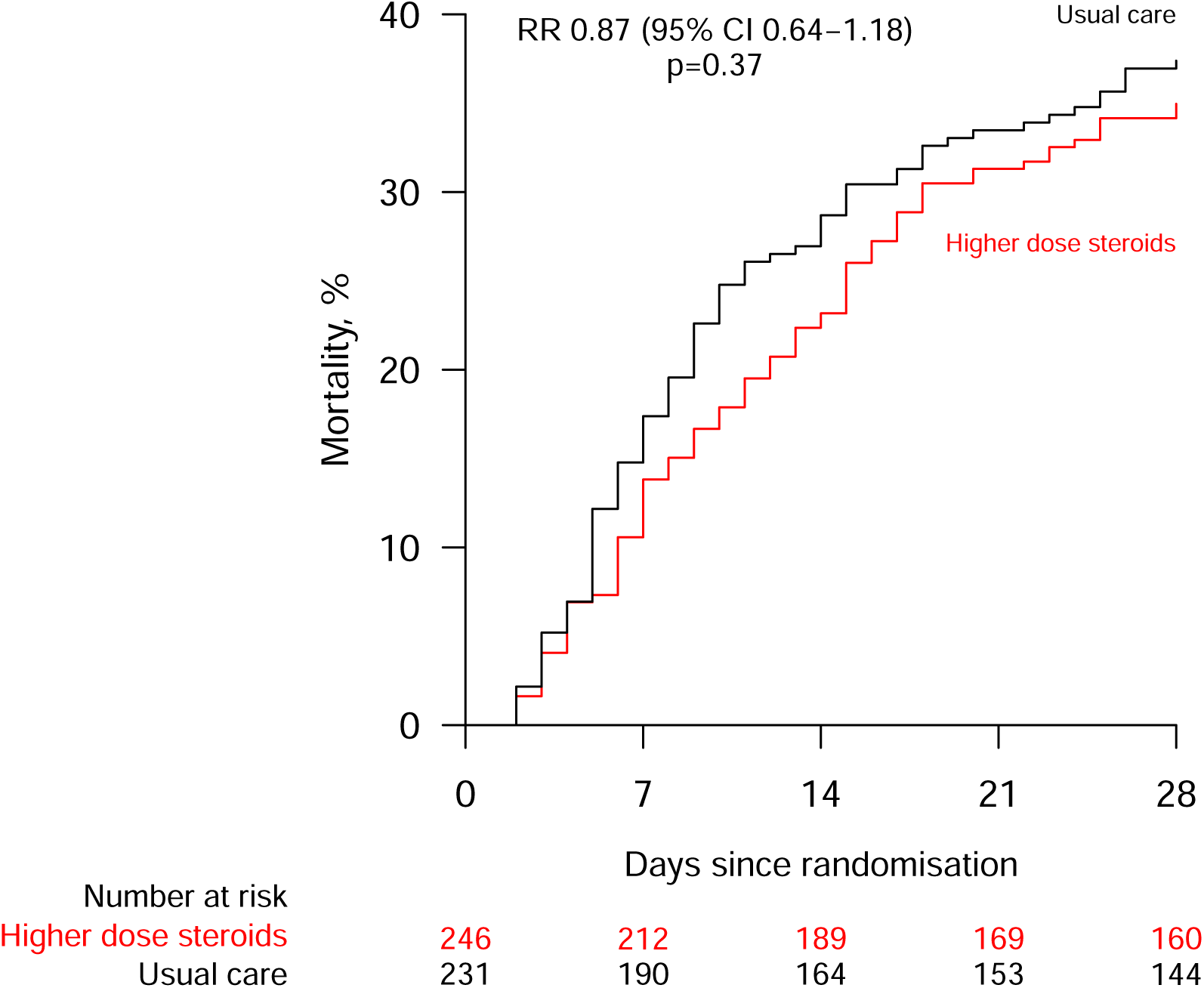
Effect of allocation to higher dose corticosteroids or usual care (lower dose corticosteroids) on 28-day mortality in patients receiving ventilatory support. RR = rate ratio

**Figure 3:**
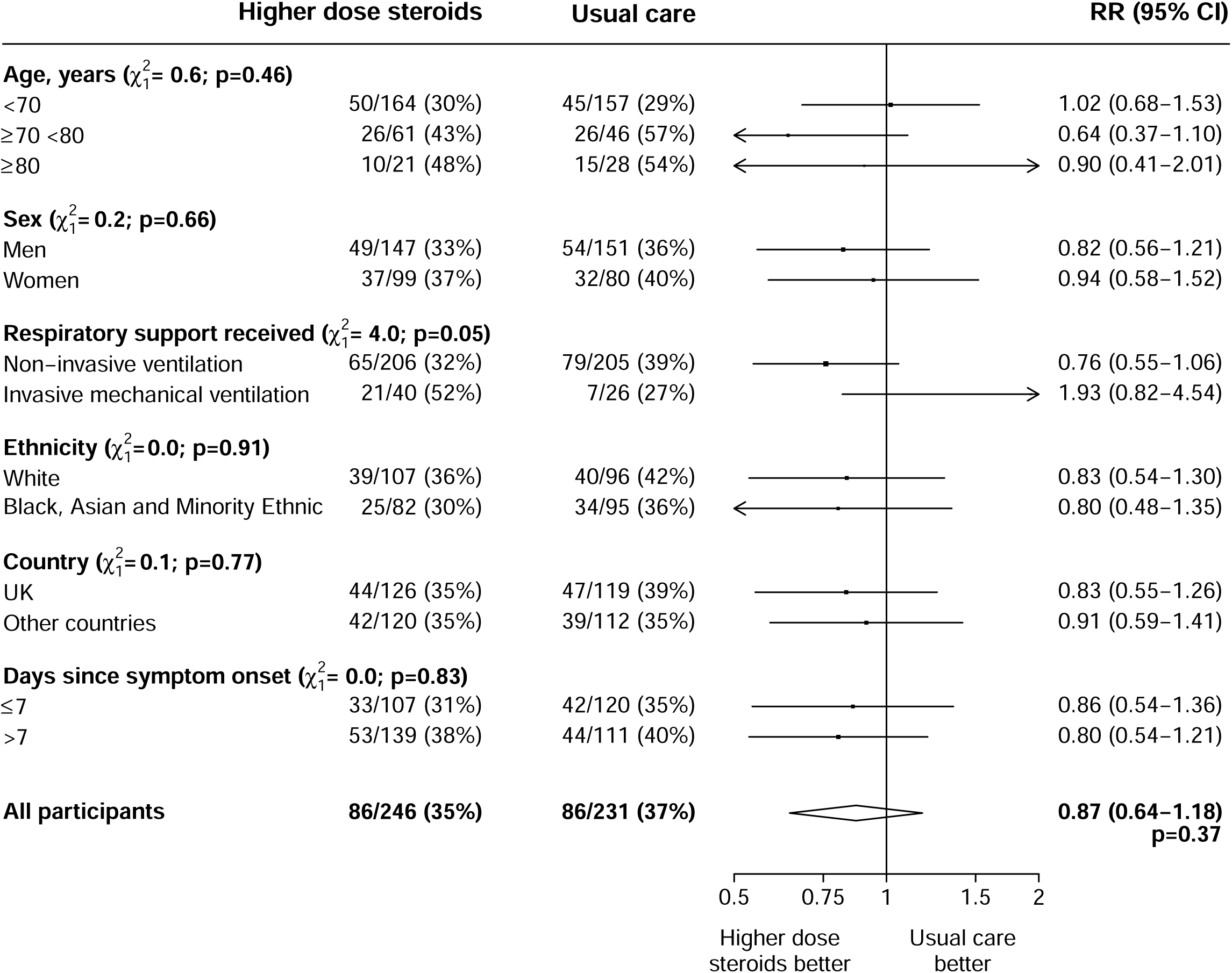
Effect of allocation to higher dose corticosteroids or usual care (lower dose corticosteroids) on 28-day mortality in patients receiving ventilatory support by other baseline characteristics. Subgroup−specific rate ratio estimates are represented by squares (with areas of the squares proportional to the amount of statistical information) and the lines through them correspond to the 95% CIs. The days since onset subgroup excludes those with missing data, but these patients are included in the overall summary diamond. RR=rate ratio.

Discharge alive within 28 days was similar among those allocated to higher dose corticosteroids compared with usual care (52% vs. 52%; rate ratio 1.04, 95% CI 0.81 to 1.33; median 24 days vs. 26 days) (table 2). Among participants not on invasive mechanical ventilation (i.e. on non-invasive ventilation) at randomisation, allocation to higher dose corticosteroids was associated with a non-significant reduction in the risk of progressing to the composite secondary outcome of invasive mechanical ventilation or death (37% vs. 45%, risk ratio 0.79, 95% CI 0.63 to 1.00) (table 2). There were no significant differences in use of invasive mechanical ventilation amongst patients on non-invasive ventilation at randomisation, or receipt of haemodialysis or haemofiltration (table 2).

137 (80%) of the deaths within 28 days were attributed to COVID-19 (webtable 3). Allocation to higher dose corticosteroids was associated with a non-significant increase in secondary infections (29% vs. 21%, absolute risk increase 7.6%, 95% CI −0.1 to 15.4%; webtable 4) including pneumonia reported as not due to COVID-19 (21% vs. 14%, absolute risk increase 6.4%, 95% CI −0.3 to 13.2%). There were no significant differences in the rates of other presentations of new onset cardiac arrhythmia, thrombotic events, clinically significant bleeding or metabolic complications (including hyperglycaemia receiving new use of insulin) (webtable 4). There were 32 reports (among 19 participants) of a serious adverse reaction believed to be related to treatment with higher dose corticosteroids (webtable 5), including 15 reports of a serious non-COVID infection, 5 with hyperglycaemia and 4 with gastro-intestinal bleeding.

Our meta-analysis identified 142 deaths among 657 participants in 5 other trials assessing higher dose corticosteroids among patients requiring ventilation (webfigure 1). Our results appeared to be consistent with the other limited data and overall there was no evidence that higher-dose corticosteroids were beneficial in these patients (overall ratio of death rates 0.87, 95% CI 0.69-1.11).

## DISCUSSION

In this randomised trial among patients with COVID-19 receiving ventilatory support, we found no evidence that allocation to higher dose corticosteroids reduced 28-day mortality, duration of hospitalization or, among patients not on invasive mechanical ventilation at baseline, the composite outcome of use of invasive mechanical ventilation or death.

The non-significant reductions observed in this population of patients contrast with the significant hazards observed among patients not receiving ventilatory support reported previously.^5^ There is statistical heterogeneity (p=0.007) between the estimates of the effect of allocation to higher dose corticosteroids on 28 day mortality in these two populations: 0.87 (95% CI 0.64-1.18) among patients receiving ventilatory supported versus 1.59 (95% CI 1.20-2.10) among those not. It is possible that the reported result among patients not receiving ventilatory support is an over-estimate of the true effect because it was based on an interim analysis, and therefore the true heterogeneity between these two populations may not be so marked.^24^ Although there is clear evidence of effect modification by baseline disease severity with low-dose corticosteroids,^1^ the subgroup analysis within the comparison reported here does not suggest that there is increasing benefit with increasing severity of disease with if anything the reverse being seen, albeit based on a very small number of outcomes.

Results from RECOVERY and other trials have shown that combining immunomodulatory therapies can provide additional benefit with both tocilizumab and baricitinib providing additional benefits over low-dose corticosteroids.^3,25^ These drugs are not widely available nor affordable in low-and middle-income countries whereas higher doses of corticosteroids are. However, we found no evidence that higher doses of corticosteroids were beneficial, even in the absence of tocilizumab, so our results do not support their use as a more affordable option where such therapies are not available.

There was a non-significant excess of secondary infections in this comparison (absolute risk increase 7.7%, 95% CI −0.1 to 15.4). The proportional increase of about one-third was similar to that observed among patients not requiring ventilatory support, so when considered in combination it is clear that higher dose corticosteroids are associated with more infections than usual care that includes low dose corticosteroids. However, the excess of hyperglycaemia requiring new use of insulin reported among patients not receiving ventilatory support was not observed in this population, but this may have been a chance finding as it is based on fewer than 100 events.

Strengths of the RECOVERY trial are that it is randomised, has a large sample size, broad eligibility criteria and more than 99% of patients in this analysis have been followed up for the primary outcome. The trial was also conducted in areas with high (south and southeast Asia, and Africa) and low (UK) prevalence of tuberculosis and other infections. The study has some limitations: this randomised trial is open label (i.e., participants and local hospital staff are aware of the assigned treatment), however, the outcome of death is unambiguous. Information on radiological, virological or physiological outcomes were not collected.

In summary, the results provide no evidence that in hypoxic COVID-19 patients receiving ventilatory support, a higher dose of corticosteroids (dexamethasone 20 mg once daily for 5 days followed by 10 mg once daily for 5 days or until discharge if sooner) provides additional benefit over low-dose corticosteroids which should remain the standard of care for such patients.

## Contributors

This manuscript was initially drafted by RH, further developed by the Writing Committee, and approved by all members of the Trial Steering Committee. PWH and MJL vouch for the data and analyses, and for the fidelity of this report to the study protocol and data analysis plan. PWH, BB, RLH, JA, JKB, MB, LCC, JD, SNF, TJ, EJ, KJ, WSL, MM, AMo, AMu, KR, GT, RH, and MJL designed the trial and study protocol. MM, LP, MC, G P-A, EK, NTP, AS, NN, RS, PNT, DP, US, RS, EN, MR and the Data Linkage team at the RECOVERY Coordinating Centre, and the Health Records and Local Clinical Centre staff listed in the appendix collected the data. NS and JRE had access to the study data and did the statistical analysis. All authors contributed to data interpretation and critical review and revision of the manuscript. PWH and MJL had access to the study data and had final responsibility for the decision to submit for publication.

## Writing Committee (on behalf of the RECOVERY Collaborative Group)

Peter W Horby*, Jonathan R Emberson*, Louise Thwaites*, Mark Campbell, Leon Peto, Guilherme Pessoa-Amorim, Natalie Staplin, Raph L Hamers, John Amuasi, Jeremy Nel, Evelyne Kestelyn, Nguyen Thanh Phong, Anil Shrestha, Nasronudin Nasronudin, Rahuldeb Sarkar, Pham Ngoc Thach, Damodar Paudel, Uun Sumardi, Richard Stewart, Erni Nelwan, Manisha Rawal, J Kenneth Baillie, Maya H Buch, Saul N Faust, Thomas Jaki, Katie Jeffery, Edmund Juszczak, Marian Knight, Wei Shen Lim, Marion Mafham, Alan Montgomery, Andrew Mumford, Kathryn Rowan, Buddha Basnyat^†^, Richard Haynes^†^, Martin J Landray^†^

* PWH, JRE and GT made an equal contribution

^+^ BB, RH and MJL made an equal contribution

## Data Monitoring Committee

Peter Sandercock, Janet Darbyshire, David DeMets, Robert Fowler, David Lalloo, Mohammed Munavvar, Adilia Warris, Janet Wittes.

## Data sharing

The protocol, consent form, statistical analysis plan, definition & derivation of clinical characteristics & outcomes, training materials, regulatory documents, and other relevant study materials are available online at www.recoverytrial.net. As described in the protocol, the Trial Steering Committee will facilitate the use of the study data and approval will not be unreasonably withheld. Deidentified participant data will be made available to bona fide researchers registered with an appropriate institution within 3 months of publication. However, the Steering Committee will need to be satisfied that any proposed publication is of high quality, honours the commitments made to the study participants in the consent documentation and ethical approvals, and is compliant with relevant legal and regulatory requirements (e.g. relating to data protection and privacy). The Steering Committee will have the right to review and comment on any draft manuscripts prior to publication. Data will be made available in line with the policy and procedures described at: https://www.ndph.ox.ac.uk/data-access. Those wishing to request access should complete the form at https://www.ndph.ox.ac.uk/files/about/data_access_enquiry_form_13_6_2019.docx and e-mailed to

## Acknowledgements

Above all, we would like to thank the thousands of patients who participated in this trial. We would also like to thank the many doctors, nurses, pharmacists, other allied health professionals, and research administrators at participating hospital organisations. Supported in the UK by staff at the National Institute of Health Research (NIHR) Clinical Research Network, NHS DigiTrials, Public Health England, Department of Health & Social Care, the Intensive Care National Audit & Research Centre, Public Health Scotland, National Records Service of Scotland, the Secure Anonymised Information Linkage (SAIL) at University of Swansea, and the NHS in England, Scotland, Wales and Northern Ireland.

The RECOVERY trial is supported by grants to the University of Oxford from UK Research and Innovation (UKRI) and NIHR (MC_PC_19056), the Wellcome Trust (Grant Ref: 222406/Z/20/Z) through the COVID-19 Therapeutics Accelerator, and by core funding provided by the NIHR Oxford Biomedical Research Centre, the Wellcome Trust, the Bill and Melinda Gates Foundation, the Foreign, Commonwealth and Development Office, Health Data Research UK, the Medical Research Council, the NIHR Health Protection Unit in Emerging and Zoonotic Infections, and NIHR Clinical Trials Unit Support Funding. TJ is supported by a grant from UK Medical Research Council (MC_UU_00002/14). WSL is supported by core funding provided by NIHR Nottingham Biomedical Research Centre. Tocilizumab, casirivimab and imdevimab, sotrovimab, and empagliflozin were provided through support from Roche, Regeneron, GSK, and Boehringer Ingelheim, respectively. Colchicine for use in Indonesia was provided by Combiphar. The views expressed in this publication are those of the authors and not necessarily those of the NHS, the NIHR or the Department of Health and Social Care. For the purpose of Open Access, the author has applied a CC BY public copyright licence to any Author Accepted Manuscript version arising from this submission.

